# Room-Aware Transformer for Activities of Daily Living Recognition in Adults with Mild Neurocognitive Disorder Using Ambient Sensors: Development and Evaluation Study

**DOI:** 10.64898/2026.11.08.26367558

**Authors:** Nader Abdalnabi, Venkatanand Ram Addepalli, Abby Roessler, Andrew Kiselica, Erich Kummerfeld, Shraboni Sarker, Praveen Rao, Knoo Lee

## Abstract

**Background:** Unobtrusive recognition of activities of daily living (ADLs) from ambient sensors may support longitudinal assessment of everyday functioning in adults with mild neurocognitive disorder (mild NCD). However, real-world activity recognition is complicated by heterogeneous home layouts, sensor configurations, routines, and class distributions.

**Objective:** This study evaluated whether a room-aware Transformer that preserves location-specific sensor structure improves participant-level ADL recognition relative to a global Transformer and a stacked unidirectional long short-term memory (LSTM) network.

**Methods:** Following pilot feasibility testing with two adult volunteers, we analyzed five homes of adults with mild NCD, comprising 92,765 minute-level observations across 67 annotated home-days. Motion, temperature, illuminance, humidity, and door-contact data were aligned to participant activity logs. The original labels were harmonized into eight categories. Models were developed separately for each home using 10-, 30-, and 60-minute windows. Complete calendar days were assigned to mutually exclusive training, validation, and chronological test sets, and no sequence crossed a day or partition boundary. The same partitions and evaluation procedures were used for all three architectures. Performance was assessed on held-out test days using accuracy, per-class F1 score, and micro- and macro-averaged one-versus-rest area under the receiver operating characteristic curve (AUC).

**Results:** The room-aware Transformer achieved the highest or tied accuracy in 9 of 15 home-window comparisons. Its accuracy ranged from 0.27 to 0.57, micro-AUC from 0.57 to 0.82, and macro-AUC from 0.54 to 0.78. Across the 15 settings, mean accuracy was similar for the room-aware and global Transformers and higher than for the LSTM. Mean micro-AUC and macro-AUC were marginally higher for the global Transformer than for the room-aware Transformer. The room-aware model showed its clearest gains in selected contexts, including Homes C and E at 10 minutes and Home A at 60 minutes, but no architecture or temporal window was uniformly optimal. Sleep/Rest was the most consistently recognized activity, whereas low-support activities frequently had low F1 scores despite moderate or high AUC values.

**Conclusions:** Room-specific feature processing provided context-dependent gains in participant-level ADL classification, particularly for accuracy in several home-window settings. The marked variation across homes, activities, and temporal windows supports personalized model development, class-sensitive evaluation, and validation in larger and more diverse residential samples before clinical use.

## Introduction

Mild neurocognitive disorder (mild NCD) is characterized by a modest decline in one or more cognitive domains beyond that expected with normal aging, while independence in everyday activities remains generally preserved [1]. Mild NCD substantially overlaps with the mild cognitive impairment (MCI), which is commonly described as an intermediate state between normal cognitive aging and dementia [2]. For some individuals, mild NCD may represent an early point along the Alzheimer’s disease and related dementias (ADRD) clinical continuum. Although substantial functional dependence is not characteristic of mild NCD, individuals may require greater effort, additional time, compensatory strategies, or accommodations to complete complex daily activities [1]. Therefore, unobtrusive recognition of activities of daily living using ambient sensors may help characterize subtle functional patterns and longitudinal changes within natural home environments.

Early detection and tracking of such functional decline is critical for timely intervention. Moreover, recognition of changes in ADL performance may aid in early diagnosis, as subtle changes in routine motion patterns, sleep–wake cycles, and household behaviors often precede clinically detected deterioration by months or even years [3]. However, traditional assessment methods rely on infrequent clinical visits or caregiver reports, which can be subjective, inconsistent, and unable to capture real-world activity patterns [5]. Low-cost sensors, such as motion, temperature, luminance, and door-contact devices, provide rich temporal signals that may capture ADL performance while preserving privacy and avoiding adherence challenges common to other methods (e.g., cameras, wearable devices) [4].

Indeed, in-home ambient sensors have gained growing attention in biomedical informatics due to their unobtrusive nature, scalability, and ability to capture minute-level activity sequences. Prior work using motion sensors, door-contact sensors, and environmental readings demonstrated potential for early detection of mild cognitive impairment (MCI) and functional change among older adults [5] [6]. Similarly, artificial intelligence (AI) models have shown promise for classifying ADLs, but many require dense instrumentation, large training sets, or computationally heavy architectures that limit practical deployment in real homes.

Deep Learning sequence models for ADL recognition may overcome some of these limitations, as they can yield high classification accuracy with lower instrumentation and sample size needs. For example, Liciotti et al reported 66.92 -94.12% accuracy for ADL recognition with LSTM-based models developed from the CASAS datasets [7]. Similarly, across five CASAS homes, Srivatsa and Plötz reported F1-scores of 0.746 to 0.901 for LSTM, 0.755 to 0.904 for transformer, and 0.783 to 0.924 for bidirectional LSTM [8]. Finally, Addepalli et al. proposed a deterministic room-specialized mixture-of-experts architecture that routed activity segments to compact room-specific transformer experts, achieving macro-F1 scores of 0.60-0.88 across five naturally occupied homes [9]. Taken together, these results suggest that deep sequence predictive models can accurately and efficiently recognize ADLs from heterogeneous sensor streams.

Of course, there were important methodological variations in how sensory data was represented spatially and temporally across these studies. Liciotti et al applied unidirectional and bidirectional LSTMs to globally represented CASAS sensor sequences, capturing temporal dependencies without explicitly preserving room-level structure [7]. Srivatsa and Plötz instead modeled sensors as a learned graph based on co-activation relationships, encoding local and global sensor dependencies without organizing features into functional room-specific branches [8]. And the room-specialized mixture of experts used deterministic hard gating to route each activity segment to a single independently trained room expert [9].

Unlike these previous studies, our investigation evaluated ADL classification in five heterogeneous, naturally occupied homes of adults with mild NCD. The proposed room-aware Transformer preserves spatial structure through dedicated room-specific feature projections followed by shared temporal self-attention, and its contribution is examined through comparisons with both a global Transformer and an LSTM baseline. Moreover, the study evaluates 10-, 30-, and 60-minute windows representing recent actions, room-transition patterns, and broader daily routines, respectively, using strictly chronological held-out complete days and both aggregate and class-sensitive performance measures.

Despite these advances, there remains a critical need for efficient, non-invasive ADL recognition systems that can operate reliably in multi-occupant households and accommodate the sparse, noisy nature of real-world sensor data. This study makes three principal contributions:

1. It introduces a room-aware Transformer that organizes ambient-sensor features by functional location and applies room-specific projections before shared temporal self-attention, thereby preserving spatial structure that global representations may obscure.
2. It evaluates the room-aware Transformer against global Transformer and stacked unidirectional LSTM baselines using 92,765 minute-level observations from five heterogeneous, real-world homes of adults with mild NCD, capturing variation in layouts, sensor configurations, activity distributions, and routines.
3. It examines 10-, 30-, and 60-minute windows representing recent actions, room transitions, and broader daily routines, respectively. The observed variation across homes indicates that temporal-context selection should account for household-specific activity patterns, rather than assume a universally optimal window.

## Methods

### Study Design

We used a two-phase design. Phase I was a feasibility and development study conducted in one home with two adult volunteers. Phase II was a real-world model development and evaluation study conducted in five instrumented homes. The two phases used the same general sensing philosophy and sequence-modeling framework but differed in participant population, household configuration and annotation ontology. For this reason, data were analyzed by phase and home, rather than pooled as if all minute-level observations were independent and interchangeable.

### Ethical Considerations

The University of Missouri Institutional Review Board reviewed and approved the study procedures (IRB #2101666). All participants, or legally authorized representatives when applicable, and participating caregivers completed the informed consent process before data collection. The sensing system did not use cameras or microphones. Sensor and activity-log data were transmitted through encrypted connections and stored on secure institutional systems, and analyses used study identifiers rather than direct personal identifiers.

### Sensing System

For both phases, this study utilized a non-wearable, in-home ambient sensing platform designed to capture continuous patterns of daily activity. The system consisted of multifunction sensors measuring motion, temperature, humidity and luminance, along with door-contact sensors capturing entry and exit events. All devices communicated through a Raspberry Pi 4 equipped with an Aeotec Z-Stick to enable local data collection and wireless connectivity. Sensors were placed in key functional areas of the homes, such as the kitchen, living room, bedroom, and bathroom, to monitor movement signals and environmental changes in light, temperature and humidity relevant to ADLs. The Raspberry Pi, running the OpenHAB home-automation framework [10], transmitted encrypted data feeds to a secure cloud server via a dedicated Wi-Fi hotspot that can be extracted from API-driven front-end.

All sensor data underwent preprocessing and synchronization procedures prior to training the deep learning models used for ADL classification. The sensing platform, developed by the University of Missouri [11], has been implemented across multiple aging-in-place research sites in Missouri.

### Study Sample and Datasets

The study datasets comprised five participant dyads enrolled in the AARG study, labeled as Home A, B, C, D and E each consisting of a participant with Mild Neurocognitive Disorder (NCD) and their informal caregiver. Participants across the five homes ranged in age from 60 to 80 years and had diagnoses of mild NCD between 2022 and 2023 and were enrolled with a clinical diagnosis of mild neurocognitive disorder. Continuous ambient sensor monitoring was conducted following enrollment, and the baseline diagnosis was used to characterize the study population. Dyads were White, non-Hispanic and lived in private homes instrumented with the ambient sensing system. Activity annotations for the participant with mild NCD were used as target labels. Homes were selected based on the availability of continuous ambient sensor streams, coupled with dense activity annotations to support the training and evaluation of deep learning models.

### Input Features

The input features used for models training were derived directly from the ambient sensing system deployed in the homes of five participants with mild NCD. The study sensing system contained three types of ambient sensors to capture heterogeneous behavioral and environmental signals. First, multifunction sensors recorded passive infrared motion events together with continuous light and temperature measurements across primary home locations. Second, the multisensor provided the same motion, light, and temperature streams while additionally capturing humidity, and were therefore exclusively placed in main bathrooms to capture humidity fluctuations indicative of showering or other hygiene-related activities. Third, contact door sensors were installed on entry points (e.g., front and garage doors) to capture open–close events associated with household transitions. Accordingly, the collected data were structured into two main types: **binary** event data (motion and door states, encoded as 0 in events of no-motion or door closure or 1 if motion detected or door open) derived from all three sensor types, and **continuous** environmental data (temperature, light, and humidity) obtained from the multifunction and multisensor devices.

Motion events were captured in an event-driven manner, whereas temperature, humidity and illuminance were reported either at fixed default intervals (temperature every 15 minutes; illuminance every 60 minutes) or when the device registered a predefined threshold change (≥1 °C for temperature; ∼200 lux for illuminance).

### Activities of Daily Living

The supervised learning target variable consisted of 13 labeled Activities of Daily Living (ADLs). To ensure clinical alignment with established functional assessment frameworks, these activities were organized into three groups. Basic ADLs were defined according to the Katz Index [12], included four ADLs: Toileting, Hygiene, Eating, and Dressing, which reflect fundamental self-care abilities essential for personal hygiene, nutrition, and autonomy. Instrumental ADLs (IADLs) followed the Lawton–Brody Scale [13] and comprised five ADLs: Medication, Cooking, Washing Dishes, Transportation (ability to travel outside the home for errands, appointments, or social activities), and Household activities. The remaining four ADLs: Sleep/Rest, Mobility (movement within house limit), Leisure/Social activity, and No Activity were categorized as Recreational/Other activities to avoid conflation with clinically validated ADL and IADL measures. No Activity was operationalized as time intervals in which no activity was logged by the participant, potentially reflecting sedentary or unreported behavior. These 13 ADL labels were derived from the participant activity logs described below and were then aligned to the time-stamped sensor data. Figure 1 visualizes a sample of one day for each home and participant with mild NCD, shown with the source labels before harmonization. The visualization highlights variations in daily routines, differences in activity distributions and recurring temporal patterns, such as sleep–wake cycles, mobility periods, and household tasks.

**Figure 1.**
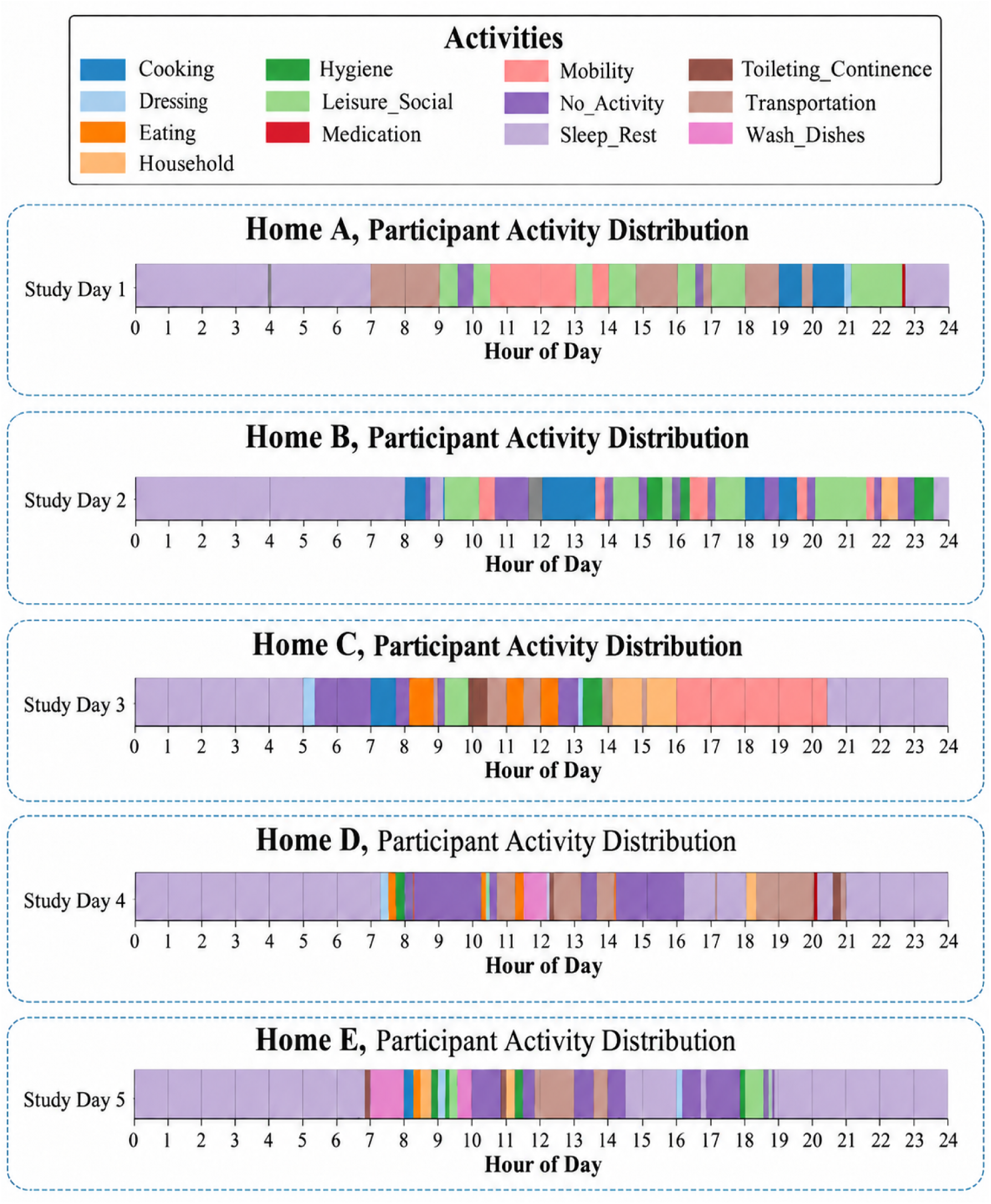
Example single-day activity timelines for participants with mild NCD in Homes A through E, shown with source labels before harmonization into the 8-category modeling ontology.

### Activity Annotation

Activity annotation for Homes A, B, C, D and E was derived from detailed activity logs completed by the participants and their informal caregivers. Activity logs captured the day date, start time, end time, activity label, location, and additional notes, enabling temporal alignment with ambient sensor streams. Both individuals received structured templates and written instructions developed by the MU research team, and informed consent was obtained prior to data collection. Participants and caregivers documented daily activities three days per week (two weekdays and one weekend day). The raw activity logs were coded and mapped to the 13 core ADLs described in the previous section. Follow-up interviews with the participants and caregivers were conducted to clarify activity patterns, validate interpretations, and refine the final target labels used for model evaluation. Final ADL target labels were aligned to a 24-hour timeline.

The final datasets comprised location-based sensor features and target labels for participants with mild NCD. Home A contained 27,046 observations and 24 features collected over 20 days, Home B contained 13,860 observations and 21 features collected over 11 days, Home C contained 23,040 observations and 15 features collected over 16 days, Home D contained 12,960 observations and 21 features collected over 9 days, and Home E contained 15,859 observations and 21 features collected over 11 days. In total, we collected 92,765 minute-level observations across 67 home-days. Differences in dimensionality reflect variations in home layouts, monitored locations, and sensor configurations, whereas differences in sample size primarily reflect differences in monitoring duration and data availability across homes.

Table 1 demonstrates substantial heterogeneity in participant activity composition across the five homes after label harmonization. Sleep/Rest was the most prevalent category in every home, ranging from 30.67% in Home A to 53.55% in Home D. Transportation accounted for 28.53% of observations in Home A, whereas Home B had greater representation of Leisure/Social (23.53%), Eating (11.66%), and Mobility (11.53%). In Home C, Sleep/Rest (46.58%) and No Activity (20.16%) together accounted for approximately two-thirds of observations. Home D had the largest proportion of Sleep/Rest (53.55%), followed by Leisure/Social (19.31%), Toileting/Continence (11.56%), and Eating (11.06%). Home E showed a more movement-oriented distribution, with Mobility accounting for 23.48% of observations. To reduce extreme sparsity and improve consistency across homes, conceptually related low-frequency activities were consolidated with functionally related categories. Specifically, Cooking was merged with Eating; Household, Washing Dishes, and Medication were merged with Leisure/Social; and Hygiene was merged with Toileting/Continence. This between-home variation supports home-specific model development and class-sensitive evaluation.

**Table 1.** Distribution of harmonized participant activity labels across five homes, n (%)

| **Activity** | **Home A<br>20 days,<br>N=27,046** | **Home B<br>11 days,<br>N=13,860** | **Home C<br>16 days,<br>N=23,040** | **Home D<br>9 days,<br>N=12,960** | **Home E<br>11 days,<br>N=15,859** |
|---|---|---|---|---|---|
| Sleep/Rest | 8,296<br>(30.67%) | 4,295<br>(30.99%) | 10,732<br>(46.58%) | 6,940<br>(53.55%) | 6,399<br>(40.35%) |
| Transportation | 7,715<br>(28.53%) | 663 (4.78%) | 2,374<br>(10.30%) | 180 (1.39%) | 1,065 (6.72%) |
| Leisure/Social | 4,480<br>(16.57%) | 3,261<br>(23.53%) | 1,815 (7.88%) | 2,502<br>(19.31%) | 3,349<br>(21.12%) |
| No Activity | 3,252<br>(12.02%) | 1,595<br>(11.51%) | 4,646<br>(20.16%) | 225 (1.74%) | 625 (3.94%) |
| Mobility | 1,251 (4.63%) | 1,598<br>(11.53%) | 1,183 (5.13%) | NA* | 3,723<br>(23.48%) |
| Eating | 895 (3.31%) | 1,616<br>(11.66%) | 1,404 (6.09%) | 1,433<br>(11.06%) | 508 (3.20%) |
| Dressing | 582 (2.15%) | NA* | 388 (1.68%) | 182 (1.40%) | 37 (0.23%) |
| Toileting/Continence | 575 (2.13%) | 832 (6.00%) | 498 (2.16%) | 1,498<br>(11.56%) | 153 (0.96%) |

(\*) NA indicates that an activity was not observed in that home.

### Deep Learning Models

The study used two temporal deep learning architectures for activity of daily living (ADL) classification from ambient sensor sequences: a stacked unidirectional Long Short-Term Memory (LSTM) network [14] and an attention-based Transformers, both global and room-aware transformers [15]. In the LSTM model, a multivariate input sequence **x**_1:*T*_ = {**x**_1_,…,**x***_T_*} was processed through one to two unidirectional LSTM layers, where hidden states were updated via gated recurrence mechanisms (input, forget, and output gates). The final hidden state served as a compact summary of temporal dynamics and was passed to a fully connected layer for ADL classification. The transformer model employed an encoder-only architecture with sinusoidal positional encodings added to the embedded inputs. Each encoder layer applied multi-head self-attention, computed as shown in eq. (1) below:

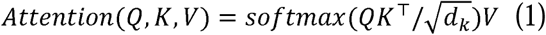

where Query (Q), Key (K), and Value (V) are learned linear projections of the input sequence embeddings. The query represents the current time step or token seeking relevant contextual information, the keys represent all time steps against which relevance is measured, and the values contain the information to be aggregated. The term 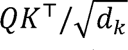 computes scaled similarity scores between queries and keys, where *d_k_* is the key dimensionality used to stabilize gradients. Applying the softmax normalizes these scores into attention weights that quantify the importance of each time step. The weighted sum of the values *V* then produces a context-aware representation that captures long-range temporal dependencies across sensor observations [15]. The core novelty of our room-aware Transformer is its hierarchical, multi-branch representation of smart-home sensor data. Rather than projecting all sensor features through a single global embedding layer, the model automatically groups features according to room prefixes, such as *bathroom_, bedroom_, kitchen_, living_ and door.* Each room group is processed by a dedicated projection block consisting of a linear layer, layer normalization, GELU activation, and dropout, allowing the model to learn representations specialized to the sensing patterns of each functional location i.e. for each room as shown in eq. (2)

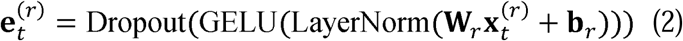

where:

- 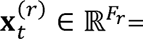 features belonging only to room *r*
- 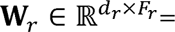 room-specific weight matrix
- *b_r_* is the corresponding bias vector usually a smaller dimension (e.g., *d/R*)

Then, temporal variables, including hour, minute, and day of the week, are processed through a separate projection block as shown in equation (3).

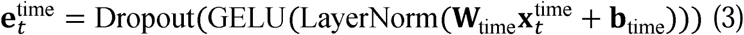

The resulting room-specific and temporal embeddings are concatenated and fused in a hidden layer before being passed to a shared temporal Transformer, which learns temporal dependencies over the fused room-level representations as shown in equation (4).

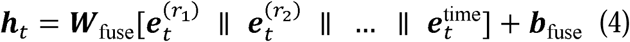

This architecture differs from a conventional global transformer by explicitly preserving room-level structure before temporal modeling; consequently, it can capture both location-specific activity signatures and home-wide temporal relationships, while accommodating differences in room availability and feature naming across homes. Figure 2 illustrates how the global and room-aware transformer architectures process and represents sensor and temporal features before shared temporal modeling and ADL classification. The key distinction between the LSTM and Transformer architectures lies in how temporal information is represented: LSTMs encode time implicitly through state propagation, whereas transformers encode it explicitly through attention over the entire sequence.

**Figure 2.**
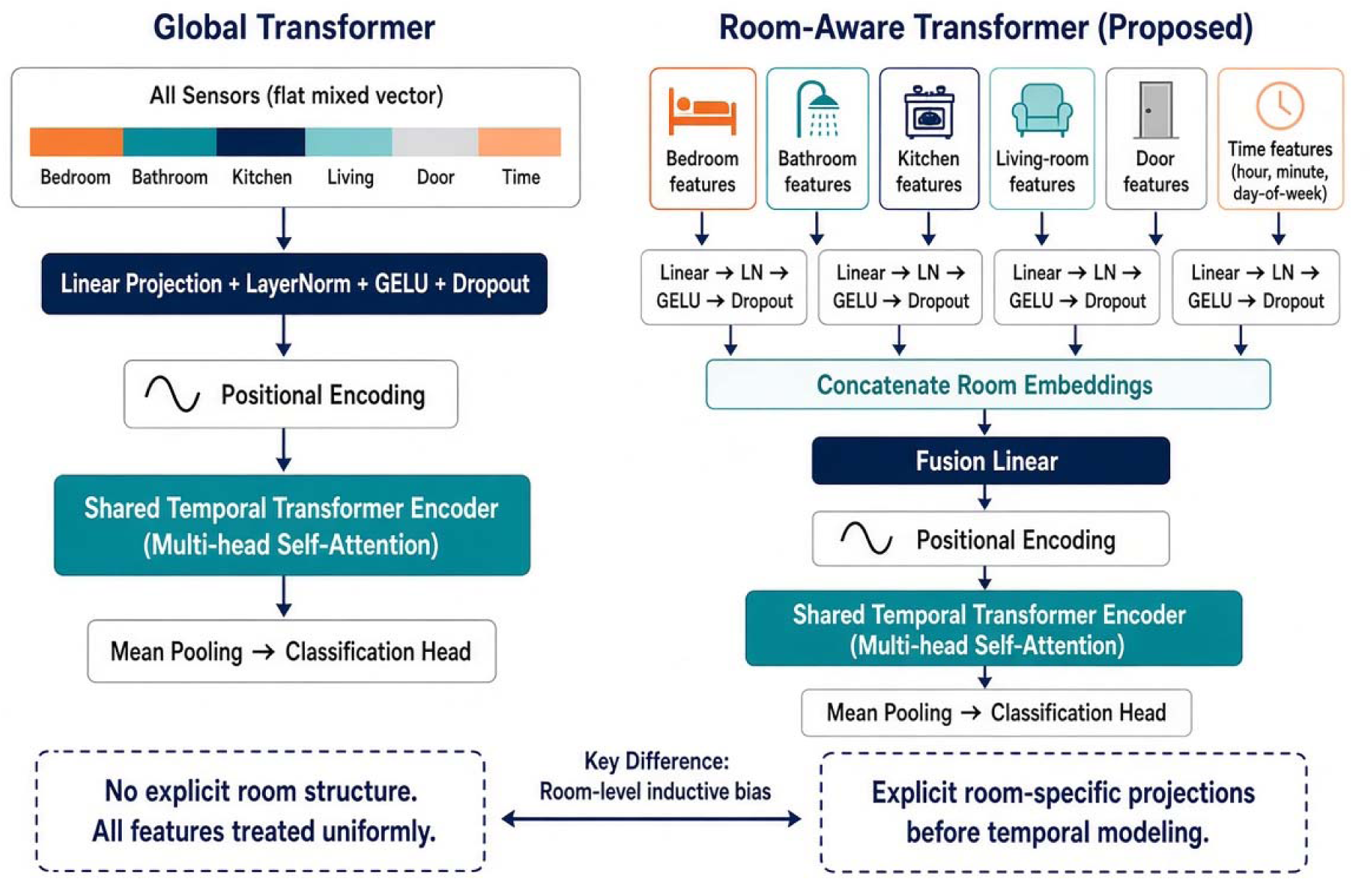
Architectural comparison of the global and proposed room-aware Transformers. LN indicates layer normalization; GELU, Gaussian error linear unit.

Figure 3 summarizes the two-phase study workflow from pilot feasibility testing to real-world evaluation. Phase I used ambient-sensor data from two adult volunteers to compare stacked LSTM and global transformer models. Phase II extended the framework to five homes of adults with mild NCD, using location-based preprocessing and the proposed room-aware transformer. Models were evaluated with 10-, 30-, and 60-minute windows using temporally partitioned training, validation, and test sets, hyperparameter tuning, and early stopping.

**Figure 3.**
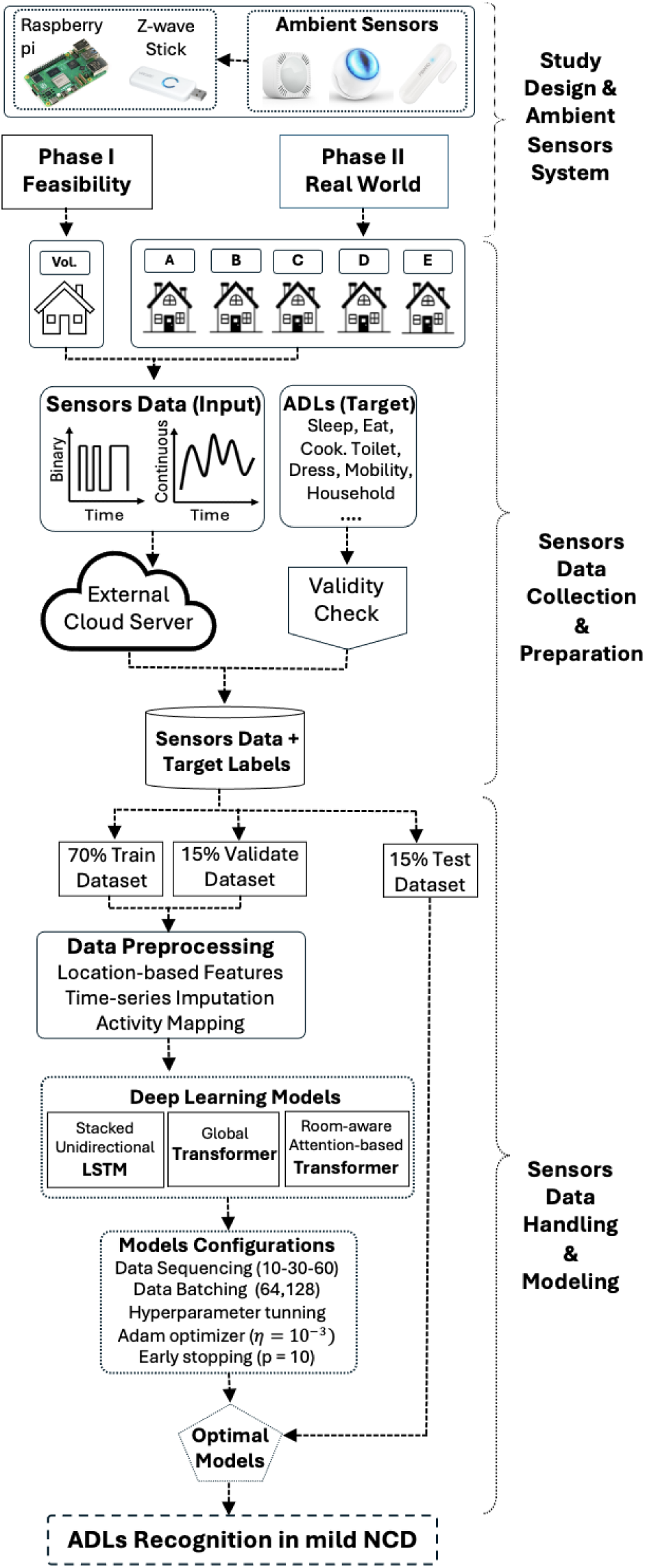
End-to-end study design and analytical workflow for ambient sensor–based activities of dailyliving (ADLs) recognition in adults with mild neurocognitive disorder (NCD)

### Data Analysis

#### Data Preprocessing

To handle the irregular time-interval readings of ambient sensors and enforce a consistent 1-minute temporal granularity, we applied two interpolation strategies. First, for continuous features (temperature, light and humidity), we used AKIMA interpolation. AKIMA [16], a locally adaptive piecewise cubic interpolation method, produced smoother trajectories while preserving the original directional trends and temporal dynamics of the sensor data. Moreover, our own pilot study found AKIMA interpolation also improved deep learning model performance when handling time-series sensors data [17]. Second, for binary event-based sensors (motion and door), missing intervals were handled using zero-filling, reflecting the absence of activity or door closure events. This strategy provides a more realistic representation of inactivity in sparse event streams and avoids biasing sequential models toward false positives (motion detected while there is no motion), ensuring temporal 1-minute consistent gap time-series across heterogeneous sensor data fed to the deep learning models.

For each home, the minute-level sensor data were partitioned at the complete-calendar-day level into mutually exclusive training, validation, and test sets. The most recent days were reserved as the chronological test set, ensuring that all test observations occurred strictly after the development period comprising the training and validation sets. This design approximated the intended prospective application, in which a model developed from previously collected sensor data is used to recognize activities on future, unseen days. Training and validation days were selected exclusively from the earlier development period, with attention to maintaining activity representation and adequate coverage of less frequent classes. All 10-, 30-, and 60-minute sequences were generated separately within their assigned days, and no sequence was permitted to cross a calendar-day or data-partition boundary. This splitting strategy is more rigorous than randomly assigning individual observations or overlapping windows because temporally adjacent samples are highly correlated and can otherwise appear in both training and testing sets, producing optimistically biased performance estimates [18]. The validation set alone guided hyperparameter selection, early stopping, and model selection, whereas the held-out test set was used only for final performance estimation, consistent with established recommendations for prediction-model development and leakage prevention [19]. Robustness was further supported through class-weighted learning, dropout, weight decay, gradient clipping, restoration of the best validation checkpoint, deterministic random seeds, and reporting of per-class F1 and macro-AUC to account for class imbalance. Importantly, identical partitions, preprocessing procedures, sequence lengths, model-selection criteria, and training settings were applied to the LSTM, Global Transformer, and Room-Aware Transformer, thereby improving the fairness and reproducibility of model comparisons. Compared with conventional random or standard k-fold splitting, this design better preserves the temporal structure of continuous in-home sensing data and provides a more realistic estimate of within-home performance on future days, although it should not be interpreted as external validation across unseen homes or participants.

#### Deep Learning Configurations

The three deep-learning architectures were trained using architecture-specific configurations appropriate to their distinct temporal-learning mechanisms, while shared experimental settings were standardized to facilitate direct comparison [14] [17] [18]. For the stacked unidirectional LSTM, input sequences of 10, 30, and 60 minutes were processed recurrently through stacked LSTM layers with predefined candidate hidden-state dimensions and layer depths. We applied dropout between recurrent layers to reduce overfitting, and the final hidden-state representation was passed to a fully connected layer for multiclass ADL classification. The global Transformer projected all sensor and temporal features jointly through a single embedding layer, added sinusoidal positional encodings, and processed the resulting representations using four encoder layers with four self-attention heads, GELU activation, feed-forward dimensions equal to four times the embedding dimension, and dropout of 0.20. In contrast, the room-aware transformer first grouped sensor features by functional-location prefixes. Each group was processed through a dedicated linear projection, layer-normalization, GELU, and dropout branch, while hour, minute, and day-of-week variables were projected separately. The room-specific and temporal embeddings were then concatenated, fused into a 16-dimensional representation, combined with sinusoidal positional encodings, and processed through the same four-layer, four-head transformer encoder used by the global architecture before temporal mean pooling and ADL classification. Across all three models, the same within-home data partitions, upstream preprocessing workflow, sequence lengths, batch-sizes of 64 and 128, class-weighted cross-entropy loss, Adam optimizer with a learning rate of 10 ³ and weight decay of 10, gradient-norm clipping at 1.0, maximum of 100 epochs, and early stopping based on validation loss with a patience of 10 epochs were applied. The same evaluation metrics were calculated for every architecture, and NumPy and PyTorch random seeds were fixed at 42 to reduce stochastic variation.

#### Model Performance Assessment

Model performances were evaluated on held-out test days using accuracy, per-class F1-score, and micro- and macro-averaged one-versus-rest area under the receiver operating characteristic curve (AUC-ROC). Accuracy quantified the overall proportion of correctly classified observations, while the per-class F1-score summarized activity-specific classification performance as the harmonic mean of precision and recall, balancing false-positive and false-negative errors. ROC-AUC was calculated across the full range of classification thresholds, rather than at a fixed threshold of 0.5, and measured the model’s ability to distinguish each ADL from competing classes [21]. Micro-AUC pooled one-versus-rest decisions across all observations and classes; consequently, it primarily reflected discrimination for prevalent activities such as Sleep/Rest, Leisure/Social, and Mobility from less prevalent activities. In contrast, macro-AUC assigned equal weight to every activity-specific AUC, ensuring that less frequent but clinically meaningful ADLs, including Toileting/Continence and Dressing, contributed equally to the overall estimate, regardless of their support.

In summary, high micro-AUC indicated good overall population-level discrimination, whereas high macro-AUC demonstrated that performance was distributed more equitably across common and underrepresented activities. Their joint interpretation prevented performance on dominant classes from obscuring weak recognition of infrequent ADLs, a well-established concern in imbalanced multiclass classification [9]. Per-class F1-score and AUC were additionally reported to identify specific activities for which favorable aggregate performance did not translate into reliable classification.

All models training and evaluation were conducted on a Linux Ubuntu 22.04 workstation running Python 3.10, PyTorch 2.5.1+cu121, and CUDA-enabled deep-learning libraries. The system was equipped with a NVIDIA GeForce RTX 4090 GPU.

## Results

### Phase I Feasibility in a Pilot Home

Phase I compared LSTM and global transformer models using data from two adult volunteers (V1 and V2). For V1, both architectures achieved similar average performance, with an accuracy of 0.72 and AUC of 0.94. For V2, the transformer improved average accuracy from 0.51 to 0.59 and AUC from 0.86 to 0.89 relative to the LSTM, with the best transformer configuration reaching an accuracy of 0.64 and AUC of 0.91. These findings established the feasibility of learning participant-specific ADL patterns from low-cost ambient sensor data and motivated Phase II, in which the approach was extended to five real-world homes involving participants with mild NCD. Phase II evaluated both the global feature representation and the proposed room-aware Transformer to accommodate differences in home layouts, monitored rooms, and activity distributions.

### Phase II Comparative Performance Across Real-World Homes and Temporal Windows

Table 2 demonstrates substantial variation in model performance across homes, evaluation metrics, and temporal-window lengths. The Room-Aware Transformer was comparatively favorable for accuracy, achieving the highest value in 8 of the 15 home-window comparisons and tying both alternative models in Home B at 10 minutes. Its largest accuracy advantages occurred in Home A at 60 minutes (0.37 vs 0.33 for the Global Transformer and 0.23 for the LSTM), Home C at 10 minutes (0.47 vs 0.42 for both alternatives), and Home E at 10 minutes (0.45 vs 0.37 and 0.34). The model also produced the highest micro-AUC across all three windows in Home D (0.80, 0.77, and 0.77) and at the 10- and 60-minute windows in Home E (0.65 and 0.62). However, its relative performance was less consistent for AUC-based discrimination. The LSTM and the Global Transformer achieved competitive performance in micro- and macro-AUC. However, Macro-AUC varied considerably, with the Room-Aware Transformer performing best in Home E at 10 and 60 minutes (0.71 and 0.67), while the Global Transformer reached 0.71 in Home B and 0.69 in Home C. Overall, room-specific feature processing was associated with more frequent improvements in accuracy than in AUC-based discrimination, and no single architecture or temporal-window length consistently performed best across all homes and metrics.

**Table 2.** ADL classification performance of LSTM (L), Global Transformer (G) and Room-aware Transformer (R) across five homes using 10-, 30-, and 60-minute temporal windows.

| Home | Window (min) | Accuracy (L/G/R) | Micro-AUC (L/G/R) | Macro-AUC (L/G/R) |
|---|---|---|---|---|
| A | 10 | 0.24 / **0.38** / 0.27 | **0.76** / 0.71 / 0.70 | **0.82** / 0.75 / 0.78 |
|  | 30 | 0.32 / 0.30 / **0.33** | **0.74** / 0.73 / 0.63 | **0.80** / 0.77 / 0.77 |
|  | 60 | 0.23 / 0.33 / **0.37** | 0.46 / 0.57 / **0.59** | **0.71** / 0.65 / 0.66 |
| B | 10 | **0.50**/ **0.50**/ **0.50** | **0.83** / 0.82 / 0.80 | 0.67 / 0.67 / **0.67** |
|  | 30 | 0.46 / 0.50 / **0.51** | **0.82** / 0.81 / **0.82** | **0.67** / 0.66 / **0.67** |
|  | 60 | 0.42 / 0.46 / **0.49** | **0.81** / 0.78 / 0.77 | 0.65 / **0.71** / 0.70 |
| C | 10 | 0.42 / 0.42 / **0.47** | 0.76 / **0.79** / 0.75 | 0.60 / **0.69** / 0.67 |
|  | 30 | **0.47** / 0.46 / 0.41 | 0.77 / **0.79** / 0.76 | 0.66 / **0.68** / 0.66 |
|  | 60 | 0.45 / **0.50** / 0.46 | 0.72 / **0.82** / 0.67 | 0.58 / **0.69** / 0.54 |
| D | 10 | 0.52 / 0.55 / **0.57** | 0.74 / 0.76 / **0.80** | 0.60 / **0.63** / 0.60 |
|  | 30 | 0.48 / 0.51 / **0.54** | 0.72 / 0.75 / **0.77** | 0.57 / **0.66** / 0.56 |
|  | 60 | **0.52** / 0.46 / 0.50 | 0.73 / 0.75 / **0.77** | 0.57 / **0.66** / 0.54 |
| E | 10 | 0.34 / 0.37 / **0.45** | 0.62 / 0.58 / **0.65** | 0.64 / 0.60 / **0.71** |
|  | 30 | 0.35 / **0.36** / 0.29 | **0.62** / 0.56 / 0.57 | **0.64** / 0.61 / 0.57 |
|  | 60 | 0.34 / **0.41** / 0.33 | 0.61 / 0.60 / **0.62** | 0.63 / 0.62 / **0.67** |

Figure 4 shows the temporal-window analysis revealed no uniform improvement as the observation period increased from 10 to 30 and 60 minutes. Shorter, 10-minute windows frequently produced the highest discrimination, particularly for the LSTM and Room-Aware Transformer. For example, the Room-Aware Transformer achieved its highest micro-AUC at 10 minutes in Homes A, D, and E (0.70, 0.80, and 0.65), while its highest macro-AUC also occurred at 10 minutes in Homes A, C, D, and E. The 30-minute window provided localized improvements, including the Room-Aware Transformer’s highest accuracy and micro-AUC in Home B (0.51 and 0.82) and the LSTM’s highest accuracy in Homes A, C, and E. In contrast, the 60-minute window was beneficial primarily for selected home-model combinations, including the Room-Aware Transformer’s highest accuracy in Home A (0.37) and the Global Transformer’s highest accuracy and micro-AUC in Home C (0.50 and 0.82). Longer windows frequently reduced AUC performance, particularly in Home A, suggesting that additional temporal context did not consistently improve activity discrimination. Overall, 10-minute windows were generally more effective for preserving short-term activity patterns, whereas the benefits of 30- and 60-minute windows were model- and home-specific.

**Figure 4.**
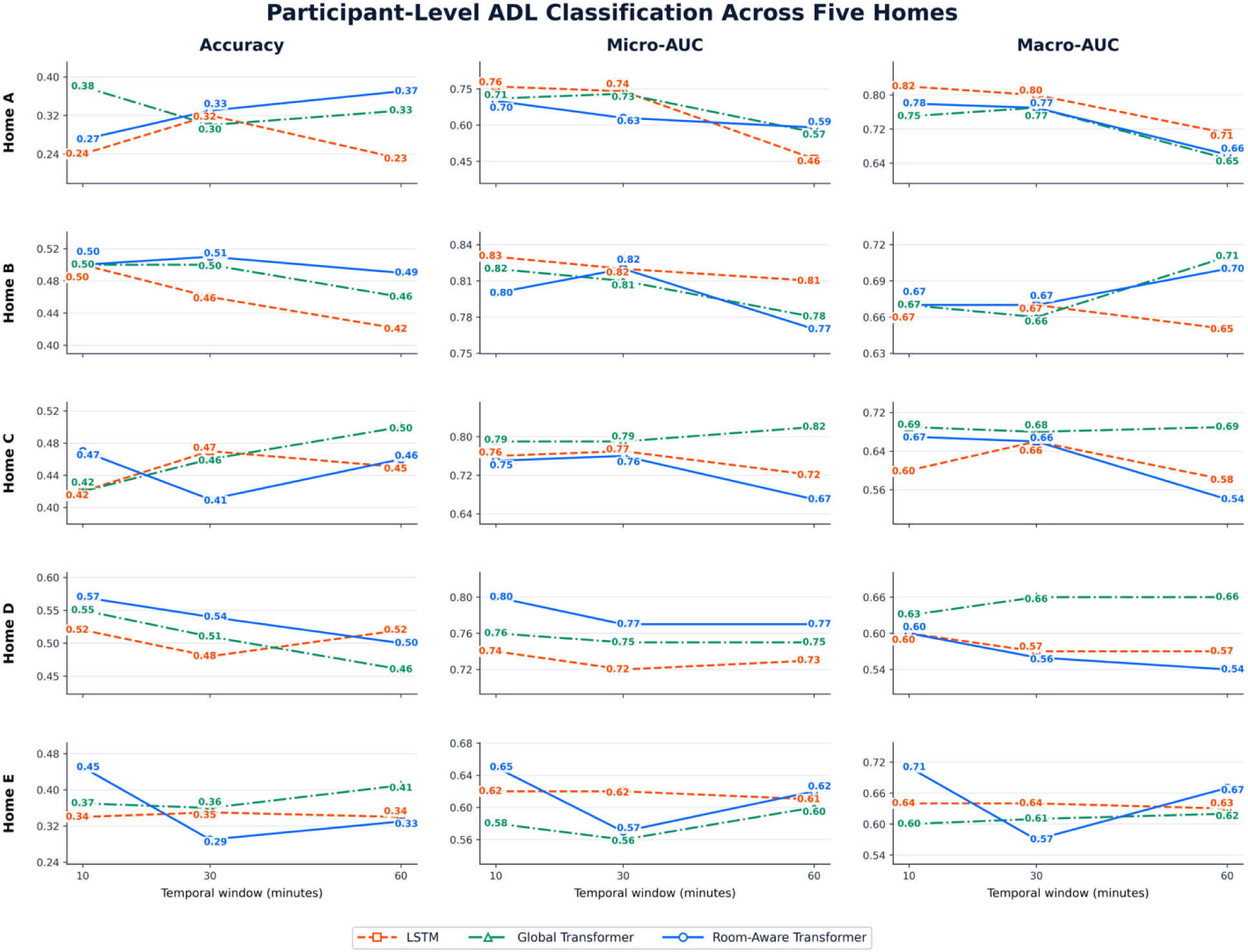
Participant-level accuracy, micro-AUC, and macro-AUC for the LSTM, global Transformer, and room-aware Transformer across five homes and 10-, 30-, and 60-minute windows. ADL indicate activity of daily living; AUC, area under the receiver operating characteristic curve; LSTM, long short-term memory.

### Activity Discrimination Across Homes

Activity-specific one-versus-rest AUC values demonstrated substantial variation in discrimination across activities, homes, and temporal windows (Figure 5). Sleep/Rest showed the most consistently high discrimination, with AUC values ranging from 0.82 to 1.00 across all homes and windows and reaching 1.00 in Home B at 60 minutes. Leisure/Social was also well distinguished in Homes A and B, with maximum AUCs of 0.80 and 0.88, respectively, while Eating demonstrated relatively stable discrimination in Homes B through D, with AUCs generally ranging from 0.67 to 0.83. Performance for Toileting/Continence, Transportation, Dressing, Mobility, and No Activity was more heterogeneous across homes, suggesting greater sensitivity to differences in activity frequency, household routines, sensor placement, and temporal context. For example, Transportation achieved an AUC of 0.85 in Home A at 10 minutes but declined with longer windows, whereas Toileting/Continence reached 0.90 in Home E at 30 minutes. At the aggregate level, the highest micro-average AUCs were observed in Home B at 30 minutes (0.82), Home B at 10 minutes (0.80), and Home D at 10 minutes (0.80). Macro-average AUC was highest in Home A at 10 minutes (0.78), indicating comparatively balanced discrimination across its represented activities. Overall, shorter windows generally provided more stable discrimination, although the optimal temporal context remained activity- and home-dependent.

**Figure 5.**
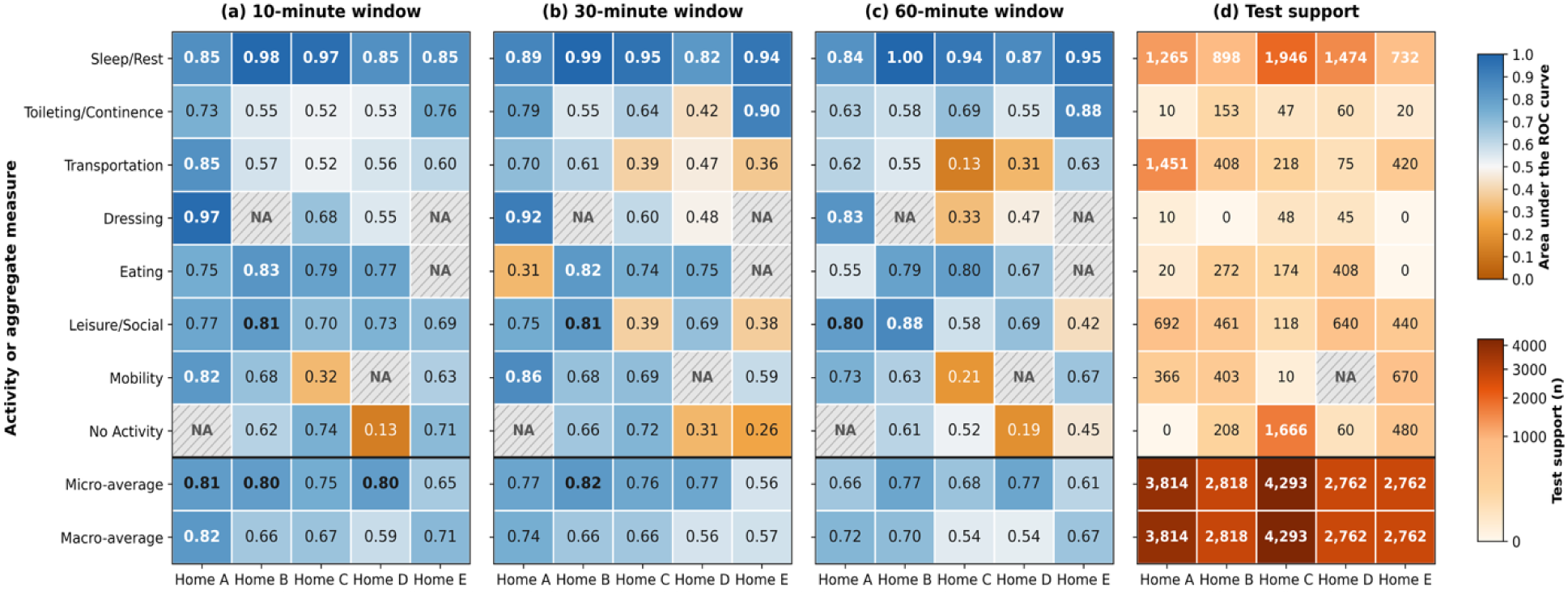
Activity-specific one-versus-rest AUC and test support for the room-aware Transformer across five homes. Panels A through C show AUC at 10-, 30-, and 60-minute windows, respectively. Panel D shows support for the 10-minute test analysis. Aggregate rows are separated from activity-specific rows. NA indicates that AUC was not estimable because the activity was absent from the corresponding test set. AUC indicates area under the receiver operating characteristic curve.

### Activity-Specific Classification Performance

Per-class evaluation of the room-aware transformer revealed substantial variation across activities, homes, and temporal-window lengths (Table 3). Sleep/Rest was the most consistently recognized activity, with F1 scores ranging from 0.47 to 0.55 in Home A, 0.89 to 0.92 in Home B, 0.81 to 0.89 in Home C, 0.78 to 0.80 in Home D, and 0.63 to 0.80 in Home E. Leisure/Social also achieved moderate performance in Homes B and D, reaching maximum F1 scores of 0.58 and 0.47, respectively, while Mobility reached 0.44 in Home A and 0.42 in Home E. Transportation was recognized most effectively in Home A at the 60-minute window (F1=0.53), but remained difficult to distinguish in the other homes. Dressing, Toileting/Continence, and several low-support activity classes generally produced low or zero F1 scores, reflecting the challenges introduced by class imbalance, limited training examples, and variation in household routines. No single temporal window consistently yielded the highest F1 score across all home-activity combinations, indicating that the amount of temporal context required for recognition depended on the activity and home environment. Overall, the findings demonstrate that the room-aware transformer captured well-represented and spatially distinctive activities more reliably than infrequent activities under real-world conditions.

**Table 3.** Per-Class F1 Scores of the Room-Aware Transformer Across Five Homes and 10-, 30-, and 60-Minute Temporal Windows

| **Activity** | **Home A\*** | **Home B** | **Home C** | **Home D** | **Home E** |
|---|---|---|---|---|---|
| Sleep/Rest | 0.55 / 0.51 / 0.47 (1225) | 0.89 / 0.92 / 0.89 (858) | 0.89 / 0.86 / 0.81 (1886) | 0.79 / 0.80 / 0.78 (1534) | 0.80 / 0.65 / 0.63 (792) |
| Dressing | 0.03 / 0.06 / 0.03 (10) | - | 0.07 / 0.00 / 0.00 (48) | 0.14 / 0.00 / 0.11 (45) | - |
| Eating | 0.00 / 0.00 / 0.00 (20) | 0.46 / 0.45 / 0.38 (272) | 0.12 / 0.00 / 0.10 (174) | 0.39 / 0.37 / 0.06 (408) | - |
| Leisure/Social | 0.28 / 0.15 / 0.35 (692) | 0.50 / 0.51 / 0.58 (461) | 0.00 / 0.00 / 0.00 (118) | 0.47 / 0.32 / 0.44 (640) | 0.15 / 0.00 / 0.00 (440) |
| Mobility | 0.07 / 0.44 / 0.35 (366) | 0.26 / 0.28 / 0.26 (403) | 0.00 / 0.00 / 0.00 (10) | - | 0.40 / 0.40 / 0.42 (670) |
| No Activity | - | 0.07 / 0.18 / 0.20 (208) | 0.29 / 0.11 / 0.38 (1666) | 0.00 / 0.00 / 0.00 (60) | 0.00 / 0.00 / 0.00 (480) |
| Toileting/ Continence | 0.00 / 0.00 / 0.01 (10) | 0.18 / 0.14 / 0.29 (153) | 0.00 / 0.03 / 0.09 (47) | 0.00 / 0.00 / 0.00 (60) | 0.35 / 0.07 / 0.00 (20) |
| Transportation | 0.35 / 0.40 / 0.53 (1431) | 0.00 / 0.26 / 0.00 (408) | 0.00 / 0.00 / 0.00 (218) | 0.00 / 0.00 / 0.00 (75) | 0.00 / 0.00 / 0.00 (420) |

(\*) Values are per-class F1 scores for the 10-, 30-, and 60-minute temporal windows, respectively. Values in parentheses represent the number of held-out test minutes (support). Support was identical across windows for most activities. A hyphen indicates that the activity was absent from the held-out test data and that its class-specific F1 score could not be estimated.

## Discussion

### Principal Findings

This two-phase study evaluated LSTM, global Transformer, and room-aware Transformer architectures for participant-level ADL classification from ambient home sensors. Phase I established feasibility in a pilot home. Phase II compared the three architectures across five homes of adults with mild NCD using identical day-level partitions and 10-, 30-, and 60-minute temporal windows. The principal finding was not uniform superiority of one architecture. Instead, the room-aware Transformer provided selective gains in accuracy in several home-window settings, while the global Transformer produced marginally higher mean micro-AUC and macro-AUC. Performance varied more by home, activity, and temporal context than the aggregate results alone would suggest.

### Comparison With Prior Work

The variation in ADL distributions across the five homes suggests that activity-recognition models must accommodate household-specific routines, rather than assume a uniform behavioral profile, consistent with prior research in ADLs recognition across different residential environments and resident populations [24]. The predominance of Sleep/Rest and sparse representation of Dressing and Toileting/Continence may favor recognition of common activities while limiting learning opportunities for underrepresented classes, reinforcing the importance of class-sensitive performance evaluation [25]. These findings motivate home-specific adaptation, supported by evidence that differences in users’ activity patterns can reduce recognition performance and that transfer learning can improve adaptation to new users [26]. However, the observed distributions should not be interpreted as evidence of differences in mild NCD severity, because annotation coverage, monitoring duration, and individual routines may also influence activity prevalence.

Across the five homes and three temporal windows, the room-aware Transformer most often achieved the highest accuracy (8 of 15 settings), although its mean accuracy matched that of the global Transformer (0.43 for both). A possible explanation for these selective gains is the room-aware architecture’s explicit separation of room-specific feature extraction from temporal modeling. Dedicated projection branches allow each room’s sensor features to be transformed independently before fusion, preserving location-specific structure that the conventional LSTM must learn implicitly from jointly presented inputs. Multi-head self-attention subsequently relates observations across the fused sequence, potentially supporting recognition of room transitions and temporally separated activity cues. This design addresses aspects of sensor and environmental heterogeneity identified as important challenges in activity recognition, although its specific contribution requires confirmation through controlled ablation studies [27], [28]. Its AUC results were mixed: it achieved the highest or tied micro-AUC in 7 settings and the highest or tied macro-AUC in 4, but its mean micro- and macro-AUC were slightly lower than those of the global Transformer, indicating that gains in some settings did not translate into uniformly better discrimination across activity classes. Moreover, the LSTM achieved the highest accuracy in Home C at 30 minutes and in Home D at 60 minutes, showing that the simpler recurrent baseline remained competitive in some settings.

The comparison of 10-, 30-, and 60-minute windows highlights an interaction between temporal context, household routines, and model architecture [29] [30]. These associations represent a design rationale rather than fixed activity durations or demonstrated class-specific optima, consistent with evidence that different activities require different amounts of historical sensor context [29]. Window effects did not follow a consistent architectural pattern: the room-aware Transformer achieved its highest accuracy at 10 minutes in Homes C, D, and E, at 30 minutes in Home B, and at 60 minutes in Home A, whereas the LSTM achieved its highest accuracy at 30 minutes in Homes A, C, and E. In principle, self-attention can directly connect temporally separated observations without passing information through successive recurrent states [15], whereas gated recurrence emphasizes local continuity; however, the present results do not show that either mechanism systematically benefited from longer windows. Thus, the findings suggest that window length should be selected on validation data for each household and architecture, rather than a universal advantage of longer sequences, consistent with prior evidence that increasing window size does not necessarily improve recognition [30].

### Clinical Implications

Shifting from model performance to downstream impact, accurate ADL classification could have implications for caregiver burden and care coordination across the ADRD continuum [31] [32]. As functional impairment progresses, caregivers experience increasing physical, psychological, social, and economic strain, with caregiver quality of life closely coupled to participant ADL dependence [31]. Anticipating ADL decline enables more timely allocation of supportive services, assistive technologies, and home-based interventions, aligning care resources with expected functional trajectories and potentially mitigating avoidable deterioration [33]. Our results offer only preliminary, model-development evidence for sensor-based activity recognition using deep learning techniques that incorporate room-specific information. Given the modest accuracy observed here, they should not be interpreted as evidence of clinical effectiveness.

Reliable classification of ADLs for aging population, regardless of cognitive diagnoses, could support independent living by transforming unobtrusive home-sensor data into longitudinal indicators of functional status and routine stability [34]. Changes in activities such as mobility, eating, medication use, hygiene, and sleep may reveal emerging functional decline before it becomes apparent during episodic clinical assessments [35] [36]. If validated, and when integrated with appropriate clinical review, ADL classification could enable timely, individualized interventions, support care planning, and help older adults remain safely at home while reducing reliance on continuous direct supervision [36].

### Limitations

This study has several limitations. First, the evaluation included only five homes with 9 to 20 annotated days per home, limiting generalizability to other residents, layouts, routines, and sensor configurations. The chronological day-level test design provides a realistic estimate of within-home performance on future days, but it is not external validation in unseen homes or participants. Second, activity labels were based on structured aligned to minute-level intervals. Despite staff review and follow-up clarification, retrospective reporting, uncertain activity boundaries, and incomplete logging may introduce label noise. Third, substantial class imbalance and the absence of some activities from individual test sets limited reliable estimation for infrequent categories. No Activity was particularly ambiguous because it could represent sedentary behavior or an unreported activity. Fourth, each home included a co-resident caregiver, and ambient sensors cannot attribute motion or environmental changes to a specific person; models therefore predicted the participant’s logged activity from household-level signals, which may add noise when the participant and caregiver were active in different rooms. Fifth, results came from a single chronological split with 2 to 3 test days per home and a single random seed, without confidence intervals, so small between-model differences should be interpreted cautiously. Finally, cognitive status was not reassessed throughout the analyzed periods, so possible changes in neurocognitive status were not modeled.

## Conclusions

This study supports the feasibility of privacy-preserving, participant-level ADL recognition from ambient sensors in homes of adults with mild NCD. The room-aware Transformer achieved favorable accuracy in selected home-window settings, particularly where spatial organization appeared informative, but it did not consistently exceed the global Transformer or LSTM across all metrics. Sleep/Rest was recognized most reliably, whereas infrequent and ambiguous activities remained challenging. The findings support further evaluation of spatially structured models using larger, more diverse samples, repeated uncertainty-aware experiments, and external validation in unseen homes. Clinical use will require evidence that reliable activity recognition translates into meaningful longitudinal assessment and decision support.

## Acknowledgments

The authors thank Sevara Pulatova and Claire Stam for their contributions to data collection.

## Data Availability

The datasets are not publicly available because they contain sensitive in-home behavioral data from participants with mild NCD. Requests may be directed to the corresponding authors and will be considered subject to IRB approval and a data use agreement.

## Authors’ Contributions

Conceptualization: NA (lead), PR and KL (equal supporting)

Data curation: NA (lead), VRA and AR (supporting)

Formal analysis: NA (lead)

Investigation: NA (lead), VRA (supporting)

Methodology: NA (lead), VRA, PR, and KL (supporting)

Project administration: NA (lead), VRA, PR and KL (supporting)

Software: NA (lead), VRA (supporting)

Supervision: PR and KL (equal supporting)

Validation: NA (lead)

Visualization: NA (lead)

Writing, original draft: NA (lead)

Writing, review and editing: NA (lead), VRA, AR, AK, EK, SS, PR, and KL (supporting)

NA conducted the primary work across the study, including study conceptualization, methodology development, data management, model implementation, formal analysis, validation, visualization, and manuscript preparation. VRA and AR contributed to data collection, and VRA also contributed to manuscript revision. PR and KL provided supervision and oversight and serve as corresponding authors.

## Abbreviations

ADL: activity of daily living
NCD: Neurocognitive disorder
ADRD: Alzheimer disease and related dementias
AI: artificial intelligence
AUC: area under the receiver operating characteristic curve
GELU: Gaussian error linear unit
LSTM: long short-term memory
MCI: mild cognitive impairment
ROC: receiver operating characteristic

## References

[1] American Psychiatric Association, Diagnostic and Statistical Manual of Mental Disorders, DSM-5-TR. American Psychiatric Association Publishing, 2022. doi: 10.1176/appi.books.9780890425787.

[2] R. C. Petersen, “Mild Cognitive Impairment,” Continuum, vol. 22, no. 2, pp. 404–418, Apr. 2016, doi: 10.1212/CON.0000000000000313.

[3] G. J. Tranah et al., “Circadian activity rhythms and risk of incident dementia and mild cognitive impairment in older women,” Ann. Neurol., vol. 70, no. 5, pp. 722–732, Nov. 2011, doi: 10.1002/ana.22468.

[4] T. L. Hayes, M. Pavel, and J. A. Kaye, “An unobtrusive in-home monitoring system for detection of key motor changes preceding cognitive decline,” in The 26th Annual International Conference of the IEEE Engineering in Medicine and Biology Society, San Francisco, CA, USA: IEEE, 2004, pp. 2480–2483. doi: 10.1109/IEMBS.2004.1403715.

[5] M. Sandhu, M. Varnfield, S. Amadoru, P. A. Yates, B. Kusy, and D. Silvera-Tawil, “Feasibility of motion sensor-based human activity recognition for supporting independence in smart homes,” Maturitas, vol. 199, p. 108632, Aug. 2025, doi: 10.1016/j.maturitas.2025.108632.

[6] I. Susnea, L. Dumitriu, M. Talmaciu, E. Pecheanu, and D. Munteanu, “Unobtrusive Monitoring the Daily Activity Routine of Elderly People Living Alone, with Low-Cost Binary Sensors,” Sensors, vol. 19, no. 10, p. 2264, May 2019, doi: 10.3390/s19102264.

[7] D. Liciotti, M. Bernardini, L. Romeo, and E. Frontoni, “A sequential deep learning application for recognising human activities in smart homes,” Neurocomputing, vol. 396, pp. 501–513, Jul. 2020, doi: 10.1016/j.neucom.2018.10.104.

[8] Srivatsa P. and T. Plötz, “Using Graphs to Perform Effective Sensor-Based Human Activity Recognition in Smart Homes,” Sensors, vol. 24, no. 12, p. 3944, Jun. 2024, doi: 10.3390/s24123944.

[9] V. R. Addepalli, P. Rao, A. Kiselica, E. Kummerfeld, N. Abdalnabi, and K. Lee, “Room-Specialized Mixture-of-Experts for In-Home ADL Recognition with Ambient Sensors,” Jun. 12, 2026, Health Informatics. doi: 10.64898/2026.06.10.26355390.

[10] S. Tsakalidis, G. Tsoulos, D. Kontaxis, and G. Athanasiadou, “Design and Implementation of a Versatile OpenHAB IoT Testbed with a Variety of Wireless Interfaces and Sensors,” Telecom, vol. 4, no. 3, pp. 597–610, Aug. 2023, doi: 10.3390/telecom4030026.

[11] K. Lee, S. Bacon, E. Conrow, E. Curtis, A. Roberts, and B. Reeder, “PRAGMATIC, SUSTAINABLE, AND COST-FEASIBLE TECHNOLOGIES TO ENABLE REMOTE CARE COORDINATION SERVICES,” Innov. Aging, vol. 7, no. Supplement_1, pp. 97–97, Dec. 2023, doi: 10.1093/geroni/igad104.0314.

[12] S. Katz, “Studies of Illness in the Aged: The Index of ADL: A Standardized Measure of Biological and Psychosocial Function,” JAMA, vol. 185, no. 12, p. 914, Sep. 1963, doi: 10.1001/jama.1963.03060120024016.

[13] M. P. Lawton and E. M. Brody, “Assessment of Older People: Self-Maintaining and Instrumental Activities of Daily Living,” The Gerontologist, vol. 9, no. 3 Part 1, pp. 179–186, Sep. 1969, doi: 10.1093/geront/9.3_Part_1.179.

[14] S. Hochreiter and J. Schmidhuber, “Long Short-Term Memory,” Neural Comput., vol. 9, no. 8, pp. 1735–1780, Nov. 1997, doi: 10.1162/neco.1997.9.8.1735.

[15] A. Vaswani, et al., “Attention Is All You Need,” Aug. 02, 2023, arXiv: arXiv:1706.03762. doi: 10.48550/arXiv.1706.03762.

[16] H. Akima, “A New Method of Interpolation and Smooth Curve Fitting Based on Local Procedures,” J. ACM, vol. 17, no. 4, pp. 589–602, Oct. 1970, doi: 10.1145/321607.321609.

[17] N. Abdalnabi et al., “Comparison of LSTM and Transformer Models for Activities of Daily Living Recognition using In-Home Ambient Sensors,” Jan. 06, 2026, Health Informatics. doi: 10.64898/2026.01.06.25342431.

[18] A. Dehghani, O. Sarbishei, T. Glatard, and E. Shihab, “A Quantitative Comparison of Overlapping and Non-Overlapping Sliding Windows for Human Activity Recognition Using Inertial Sensors,” Sensors, vol. 19, no. 22, p. 5026, Nov. 2019, doi: 10.3390/s19225026.

[19] S. E. Davis, M. E. Matheny, S. Balu, and M. P. Sendak, “A framework for understanding label leakage in machine learning for health care,” J. Am. Med. Inform. Assoc., vol. 31, no. 1, pp. 274–280, Dec. 2023, doi: 10.1093/jamia/ocad178.

[20] F. Ordóñez and D. Roggen, “Deep Convolutional and LSTM Recurrent Neural Networks for Multimodal Wearable Activity Recognition,” Sensors, vol. 16, no. 1, p. 115, Jan. 2016, doi: 10.3390/s16010115.

[21] T. Fawcett, “An introduction to ROC analysis,” Pattern Recognit. Lett., vol. 27, no. 8, pp. 861–874, Jun. 2006, doi: 10.1016/j.patrec.2005.10.010.

[22] N. Abdalnabi et al., “Comparison of LSTM and Transformer Models for Activities of Daily Living Recognition using In-Home Ambient Sensors,” Jan. 06, 2026, Health Informatics. doi: 10.64898/2026.01.06.25342431.

[23] D. J. Cook, A. S. Crandall, B. L. Thomas, and N. C. Krishnan, “CASAS: A Smart Home in a Box,” Computer, vol. 46, no. 7, Jul. 2013, doi: 10.1109/MC.2012.328.

[24] D. Cook, “Learning Setting-Generalized Activity Models for Smart Spaces,” IEEE Intell. Syst., vol. 27, no. 1, pp. 32–38, Jan. 2012, doi: 10.1109/MIS.2010.112.

[25] Haibo He and E. A. Garcia, “Learning from Imbalanced Data,” IEEE Trans. Knowl. Data Eng., vol. 21, no. 9, pp. 1263–1284, Sep. 2009, doi: 10.1109/TKDE.2008.239.

[26] S. An, G. Bhat, S. Gumussoy, and U. Ogras, “Transfer Learning for Human Activity Recognition using Representational Analysis of Neural Networks,” Feb. 23, 2021, arXiv: arXiv:2012.04479. doi: 10.48550/arXiv.2012.04479.

[27] X. Ye, K. Sakurai, N.-K. C. Nair, and K. I.-K. Wang, “Machine Learning Techniques for Sensor-Based Human Activity Recognition with Data Heterogeneity—A Review,” Sensors, vol. 24, no. 24, p. 7975, Dec. 2024, doi: 10.3390/s24247975.

[28] K. Chen, D. Zhang, L. Yao, B. Guo, Z. Yu, and Y. Liu, “Deep Learning for Sensor-based Human Activity Recognition: Overview, Challenges, and Opportunities,” ACM Comput. Surv., vol. 54, no. 4, pp. 1–40, May 2022, doi: 10.1145/3447744.

[29] N. C. Krishnan and D. J. Cook, “Activity recognition on streaming sensor data,” Pervasive Mob. Comput., vol. 10, pp. 138–154, Feb. 2014, doi: 10.1016/j.pmcj.2012.07.003.

[30] O. Banos, J.-M. Galvez, M. Damas, H. Pomares, and I. Rojas, “Window Size Impact in Human Activity Recognition,” Sensors, vol. 14, no. 4, pp. 6474–6499, Apr. 2014, doi: 10.3390/s140406474.

[31] S.-T. Cheng, “Dementia Caregiver Burden: a Research Update and Critical Analysis,” Curr. Psychiatry Rep., vol. 19, no. 9, p. 64, Aug. 2017, doi: 10.1007/s11920-017-0818-2.

[32] M. Gómez-Gallego, J. Gómez-Amor, and J. Gómez-García, “Determinants of quality of life in Alzheimer’s disease: perspective of patients, informal caregivers, and professional caregivers,” Int. Psychogeriatr., vol. 24, no. 11, pp. 1805–1815, Nov. 2012, doi: 10.1017/S1041610212001081.

[33] L. A. Jennings et al., “Health Care Utilization and Cost Outcomes of a Comprehensive Dementia Care Program for Medicare Beneficiaries,” JAMA Intern. Med., vol. 179, no. 2, pp. 161–166, Feb. 2019, doi: 10.1001/jamainternmed.2018.5579.

[34] A. Pérez-Vereda, J. Fontecha, A. Sanchez-Miguel, L. Cabañero, I. González, and C. Nugent, “Activities of Daily Living Detection through Energy Consumption Data and Machine Learning to Support Independent Aging,” J. Med. Syst., vol. 49, no. 1, p. 124, Oct. 2025, doi: 10.1007/s10916-025-02256-2.

[35] M. E. Mlinac and M. C. Feng, “Assessment of Activities of Daily Living, Self-Care, and Independence,” Arch. Clin. Neuropsychol. Off. J. Natl. Acad. Neuropsychol., vol. 31, no. 6, pp. 506–516, Sep. 2016, doi: 10.1093/arclin/acw049.

[36] J. A. Kaye et al., “Intelligent Systems For Assessing Aging Changes: home-based, unobtrusive, and continuous assessment of aging,” J. Gerontol. B. Psychol. Sci. Soc. Sci., vol. 66 Suppl 1, no. Suppl 1, pp. i180–190, Jul. 2011, doi: 10.1093/geronb/gbq095.

